# Mode Connections For Clinical Incremental Learning: Lessons From The COVID-19 Pandemic

**DOI:** 10.1101/2023.05.05.23289583

**Authors:** Anshul Thakur, Chenyang Wang, Taha Ceritli, David Clifton, David Eyre

## Abstract

Dynamic distribution shifts caused by evolving diseases and demographic changes require domain-incremental adaptation of clinical deep learning models. However, this process of adaptation is often accompanied by *catastrophic forgetting*, and even the most sophisticated methods are not good enough for clinical applications. This paper studies incremental learning from the perspective of *mode connections*, that is, the low-loss paths connecting the minimisers of neural architectures (modes or trained weights) in the parameter space. The paper argues for learning the low-loss paths originating from an existing mode and exploring the learned paths to find an acceptable mode for the new domain. The learned paths, and hence the new domain mode, are a *function* of the existing mode. As a result, unlike traditional incremental learning, the proposed approach is able to exploit information from a deployed model without changing its weights. Pre-COVID and COVID-19 data collected in Oxford University hospitals are used as a case study to demonstrate the need for domain-incremental learning and the advantages of the proposed approach.

## 1 Introduction

Healthcare is poised to undergo a revolution as deep learning is set to provide improved diagnostics (Liu et al., 2019), personalised medicine (Wilkinson et al., 2020), efficient critical care (Sun et al., 2019) and effective management of critical resources (Rajkomar et al., 2018). Despite its advantages, deep learning models are susceptible to distribution shifts and may exhibit a noticeable performance drop if there is a shift in training and test data distributions. These distributional shifts are quite common in healthcare as diseases, treatment regimes and demographics constantly evolve (Thakur et al., 2023). Consequently, the deployed clinical models may become ineffective over time. Domain-incremental learning, a form of continual learning, allows the trained/deployed models to adapt to new domains (characterised by distribution shifts) while retaining the previously acquired information (Chen & Liu, 2018; Lee & Lee, 2020). As a result, the updated model can be employed on the new as well as the previously seen domains. The test examples used in many clinical applications can be taken from previous domains. For instance, distribution shifts caused by the COVID-19 pandemic might be reversed after the pandemic. However, domain-incremental methods are not perfect and often exhibit a performance loss on the previous domains after updating the model (Armstrong & Clifton, 2021). Thus, these methods are not suitable for sensitive clinical applications.

This paper studies incremental learning from a new perspective of *mode connectivity* and conceptualises an incremental learning framework as an elaborate network of modes in the parameter space. A mode connection is defined as the low-loss path between two minimisers of a neural network, referred to as *modes* (minima or trained weights for a task in parameter space), such that every point on this path provides either equivalent or lower loss than minimisers at the end points (Tatro et al., 2020; Garipov et al., 2018).

Deviating from the earlier work, we extend the concept of mode connectivity to learn a non-linear low-loss path for the new domain between the *existing mode* (a currently deployed model) and a *randomly chosen point* in the parameter space. This learned path is analysed to find the most suitable possible mode for the new domain, and the section of path between the existing mode and the newly identified mode is incorporated into the incremental mode network (Figure 1). As the semantics of the earlier and new domains are expected to be the same, the section of the path near the existing mode exhibits a lower loss than the section of the path near the random point. Consequently, we find the new domain mode near the existing mode and every point along the path between these modes is expected to be a viable minimiser. Thus, the proposed approach is able to expand on the existing domain knowledge, represented by the deployed model, without altering its trained weights and hence, alleviating the possibility of catastrophic forgetting.

**Figure 1:**
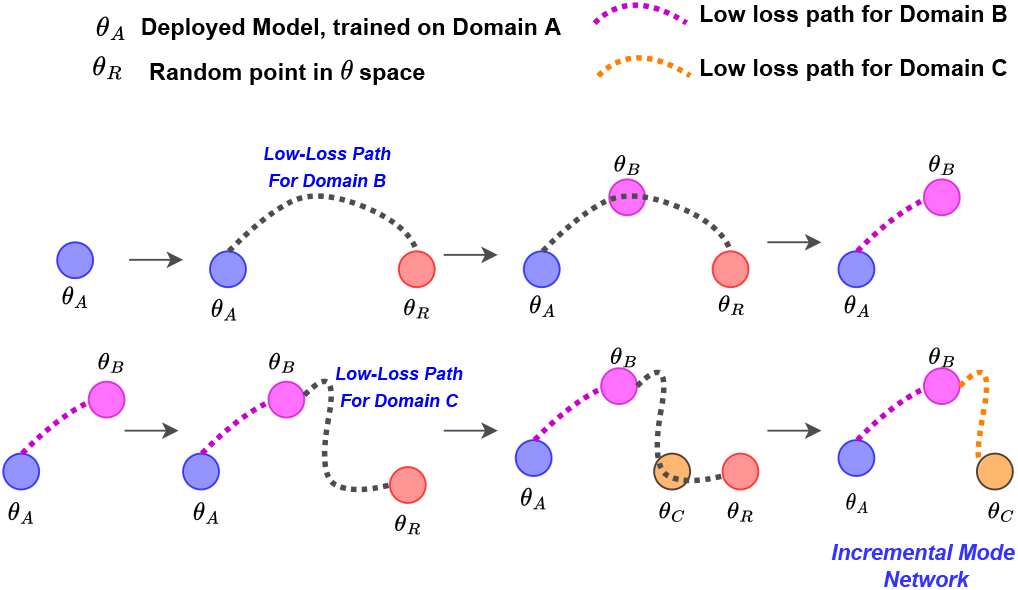
An illustration of the proposed incremental learning approach. Given a model trained on domain A, a network of modes is learned for incorporating domain B and C in an incremental manner.

The main contribution of this paper are:

- The paper conceptualises incremental or continual learning as an incremental network of modes that avoids catastrophic forgetting.
- The proposed framework results in an entire path of possible minimisers for the new domain. The modes sampled from these low-loss paths can be used as an ensemble and hence, obtain predictions with less uncertainty.
- This paper provides evidence in favour of the proposed framework by utilising the Oxford University Hospitals (OUH) data, collected between 2019 and 2021.

## 2 Proposed method

### 2.1 Mode Connections

Suppose *θ*_1_ ∈ *R*^*N*^ and *θ*_2_ ∈ *R*^*N*^ be the two modes of a neural network trained for any particular task. Then, the mode connection 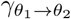 is defined as a path (mostly non-linear) between *θ*_1_ and *θ*_2_ such that every point on this path is an *effective minima* for the task. As we traverse from one end-point to another on the path, we do not witness any *loss barrier* i.e. the increment in the training loss. 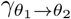 can be obtained by training any non-linear parametric curves such as *Bezier curve*.

Bezier curve *ϕ*_*θ*_(*t*), parameterised by *θ* ∈ *R*^*N*^, can be used to learn 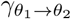 as Garipov et al. (2018):

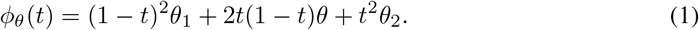

Here, *t* is the input to *ϕ*_*θ*_ and its value decides the location on the curve. *t* = 0 results in *θ*_1_ and *t* = 1 outputs *θ*_2_. Hence, the range of *t* is [0, 1].

As described in Garipov et al. (2018), *ϕ*_*θ*_ is trained by updating *θ* to achieve minimum loss at different values of *t*^1^, and hence, obtaining the desired 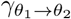. The computational effort required to learn 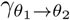 is equivalent to only training a neural network.

### 2.2 Mode Network for incremental learning

As described in Section 1, the proposed *mode network* starts with one mode (*θ*_*A*_) representing the currently deployed model. To incorporate a new mode *θ*_*B*_ (corresponding to new domain) to the mode network, we learn a mode connection 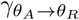 between *θ*_*A*_ and a random point in parameter space, *θ*_*R*_, for the new domain data using Bezier curve *ϕ*_*θ*_ defined in equation 1.

We identify *θ*_*B*_ as the point on the trained *ϕ*_*θ*_ that provides minimum loss for validation data 𝒟_VAL_ of the new domain:

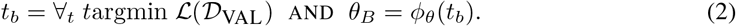

This *θ*_*B*_ and 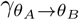 or *ϕ*_*θ*_ with truncated domain (0, *t*_*b*_] is added to the mode network (as illustrated in Figure 1). More modes corresponding to new domains can be added to the mode network by following the same procedure with the latest added mode as the starting point.

The modes or parameters are sampled from the trained mode network as per the domain of an input example. We can sample either the identified modes (such as *θ*_*B*_) or we can sample multiple modes from the low-loss paths (such as 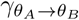) and use them as an ensemble to obtain the prediction. This work only deals with the former case. The domain of a test example can be identified using the simple domain identification methods as the one described in Section C of the appendix.

## 3 Experiments and Results

### 3.1 Task and Dataset

The proposed incremental framework is trained for the task of respiratory deterioration prediction that deals with predicting the escalation in the level of oxygen support requirements or an unplanned ICU admission (as a proxy for mechanical ventilation) in the next 24 hours (Section A.1 of the appendix) (Youssef et al., 2020).

Patient records from the *Infections in Oxfordshire Research Database* (IORD) are used for performance evaluation. The data collected from patients admitted to Oxford University hospitals (OUH) between January 2019 and June 2021 is used. Patients admitted in 2019 exhibited various underlying conditions such as pneumonia, heart failure, and asthma. In contrast, the data between March 2020 and June 2021 is only collected from patients with PCR-confirmed COVID-19. To simulate an incremental learning setup, we temporally divide the data into three subsets: *2019 dataset*, the first COVID-19 dataset (March 2020 to July 2020), and the second COVID-19 dataset (August 2020 to June 2021) dataset. The first COVID-19 dataset (*COVID-1*) corresponds to the first COVID-19 wave, whereas the second COVID-19 dataset (*COVID-2*) corresponds to the second and third waves.

Each example is represented by a 77-dimensional feature vector and a binary label (retrospectively generated) signifying respiratory deterioration within the next 24 hours. Features include demographic characteristics, vital sign measurements, laboratory test results, and inspired oxygen concentration (FiO_2_). More details about the datasets, pre-processing and feature representation are presented in Section A of the appendix. A three-layered dense neural network (Section B) is used for predicting respiratory deterioration based on the 77-d features.

### 3.2 Experiments

The first experiment is designed to analyse the predictive capabilities of the modes and the mode connections obtained using the proposed method. The model trained on the 2019 dataset is used as the starting point to learn the modes for the *COVID-1* and *COVID-2* datasets. The performance of these modes is compared against the fine-tuning or transfer learning. *2019* model is fine-tuned or adapted using *COVID-1* data and the resultant model is further adapted using *COVID-2* datasets.

The second experiment is designed to evaluate the trained mode network as a full-fledged incremental learning framework. We compare the performance of the proposed approach against GDumb (Prabhu et al., 2020) and gradient episodic memory (Lopez-Paz & Ranzato, 2017). In this experiment, we compare the performance on the new (*COVID*) as well as the previously seen domains (*2019*).

### 3.3 Results & Discussion

Figure 2 depicts the validation performance of the *mode connections* or low-loss paths learned between **(A)** *2019* mode and a random point and **(B)** *COVID-1* mode and a random point on *COVID-1* and *COVID-2* datasets, respectively. The analysis of this figure shows that the proposed approach of extending mode networks or mode connections from an *origin mode* can indeed result in effective modes for the new domains. On moving away from *t* = 0 (existing mode), we witness a significant performance boost till the loss barrier is encountered near the random end point (*t* = 1).

**Figure 2:**
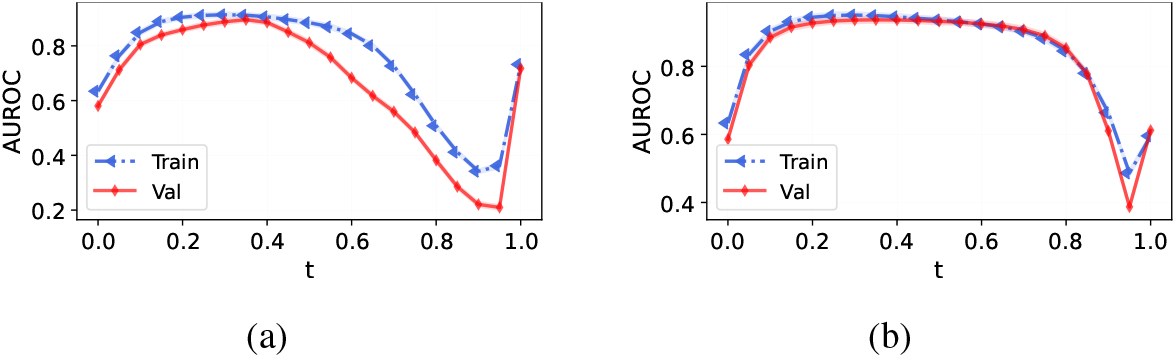
Performance of different points on trained Bezier curve or mode connections originating from (a) *2019* and *COVID-1* mode for the *COVID-1* and *COVID-2* datasets, respectively.

Table 1 tabulates the performance of the learned modes against the comparative methods. The analysis of this table shows that the performance of the proposed method is either comparable or better than the transfer learning. Hence, similar to transfer learning, the learned modes are indeed exploiting the information present in the origin modes. However, unlike transfer learning, these modes are obtained without altering the origin or previous domain models. Another important highlight of this table is the failure of *2019* model on both COVID datasets. This shows that there are huge distribution shifts between the pre-COVID and COVID data (Figure 3 of the appendix) that rendered the pre-COVID model nearly useless on COVID-19 patients.

**Table 1:**
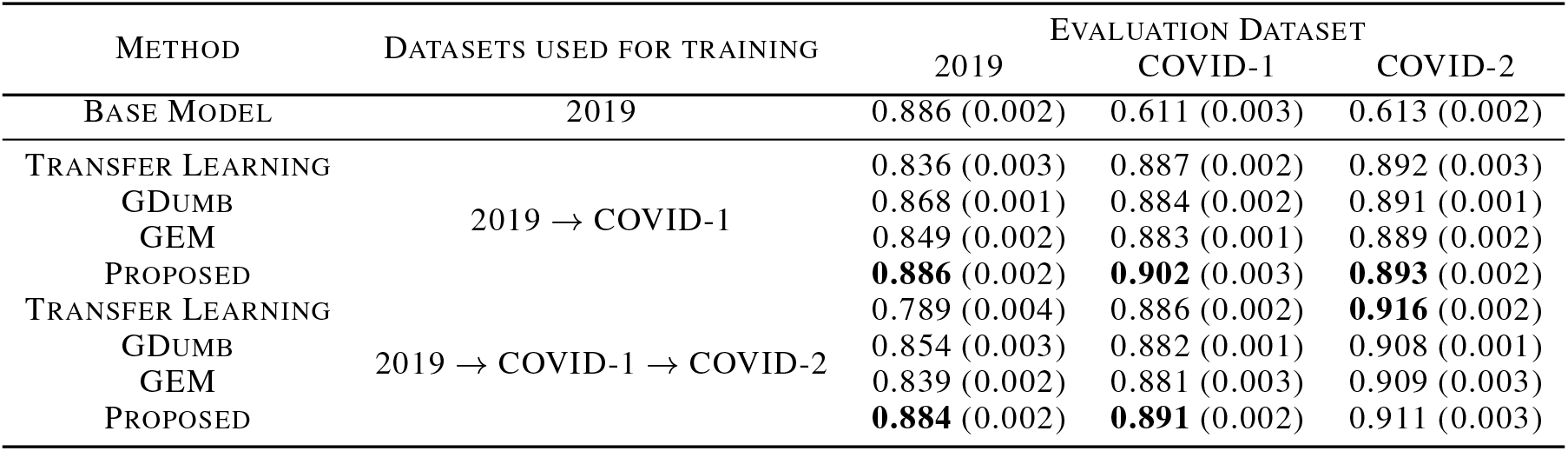
Performance of the proposed mode network against the comparative methods. AUROC is used as the performance metric.

**Figure 3:**
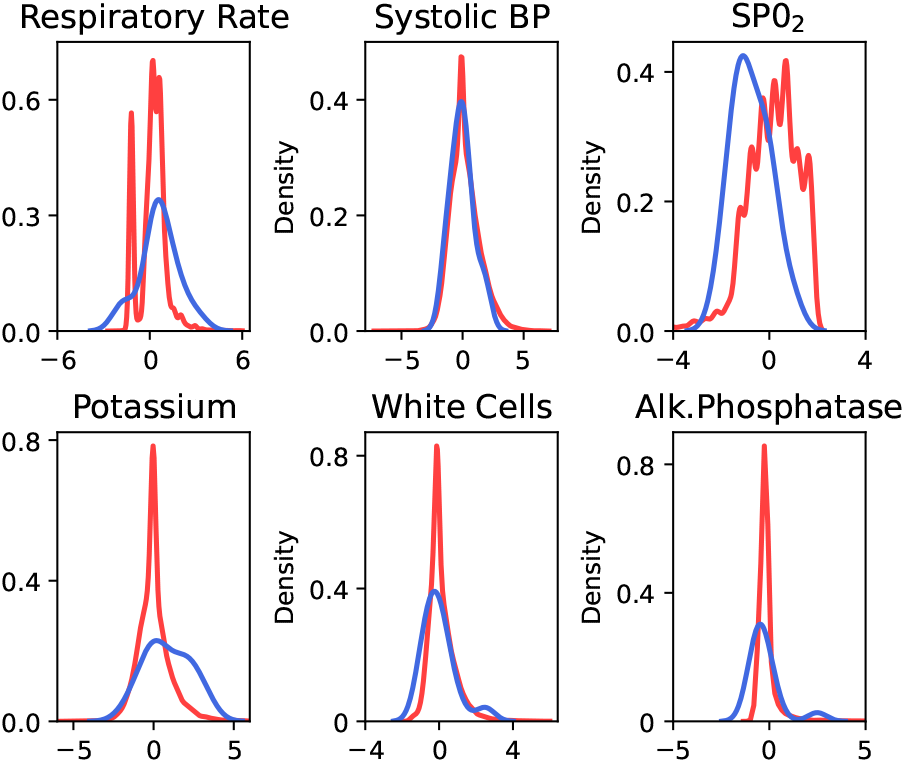
Kernel density estimation (KDE) plots highlighting distribution shifts between 2019 and COVID-1 datasets for 6 randomly chosen features.

Table 1 further documents the performance comparison of the proposed method against the comparative incremental learning methods. The analysis of this table highlights that the proposed framework exhibits no catastrophic forgetting while effectively performing on the newly introduced domains. This shows that the proposed approach results in a mode network that acts as an effective continual or incremental learning framework.

## 4 Conclusion

This paper presented a mode connection-based approach for performing incremental learning. The proposed method doesn’t suffer from catastrophic forgetting while learning new modes or domains. Experimental results on the OUH data highlighted the potential benefits of the proposed method. Although this paper presented strong evidence in favour of the mode networks as incremental learning frameworks, a thorough investigation is required to analyse the behaviour of this method in more challenging and diverse conditions. More work is required to bring down the computational and space complexity of the proposed method. Moreover, we did not emphasise the ensembling aspect of the mode connections that has been used for bringing down the uncertainty in predictions. Future work will deal with these aspects to refine the proposed approach.

## Data Availability

All data is publicly available

## A Data details

### A.1 Respiratory Support Levels

The level of respiratory support is defined based on the oxygen delivery devices. A list of the devices and the corresponding level of support is documented in Table 2.

**Table 2:** Different respiratory support levels and the corresponding oxygen support devices used at each level.

| SUPPORT LEVEL | DEVICES |
| --- | --- |
| LEVEL 0 | NO SUPPORT REQUIRED |
| LEVEL 1 | SIMPLE MASK, VENTURI FACE MASK, NASAL CANNULAE, NEBULISER MASK, OXYMASK, TRACHEOSTOMY MASK |
| LEVEL 2 | RESERVOIR MASK, NON-REBREATHER MASK |
| LEVEL 3 | HIGH FLOW, NON-INVASIVE SYSTEM, CONTINUOUS POSITIVE AIRWAY PRESSURE (CPAP), AIRVO |

### A.2 Patient Characteristics

Table 3 documents the characteristics of patients used in this study.

**Table 3:**
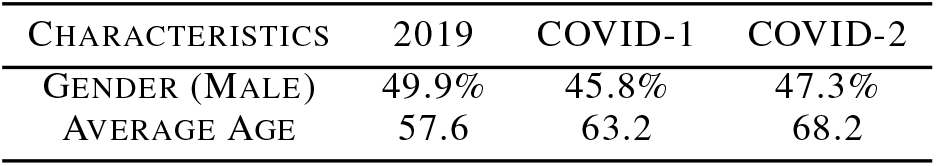
Patient characteristics in the used data.

| CHARACTERISTICS | 2019 | COVID-1 | COVID-2 |
| --- | --- | --- | --- |
| GENDER (MALE) | 49.9% | 45.8% | 47.3% |
| AVERAGE AGE | 57.6 | 63.2 | 68.2 |

### A.3 Pre-processing

Data collected between December 2019 - February 2020 is removed to prevent potential data contamination from undiagnosed COVID-19 cases. For patients with multiple respiratory deterioration events, we removed observations recorded after the first event. Implausible physiological values were excluded, and non-numerical readings were replaced with clinically appropriate values. If a lab reading is below or above the detection threshold of the laboratory assay, the reading is replaced with zero or an appropriate value to maintain the significance of the high result, respectively. If no information is available regarding the provision of supplemental oxygen, it is assumed that no supplemental oxygen was provided. Similarly, the same assumption is made for AVPU, and missing AVPU values are replaced with ‘alert’. For other missing values, we have used median-based imputation.

### A.4 Number of patients in each sub-dataset

Table 4 documents the number of patients, number of samples, and percentage of samples exhibiting respiratory deterioration in each sub-dataset. We divided each dataset into 65% training, 15% validation and 20% test sets.

**Table 4:** Number of patients and examples in each sub-dataset obtained from Oxford University hospitals (OUH) data.

| DATASET | # PATIENTS | # EXAMPLES | # RESPIRATORY DETERIORATION EVENTS |
| --- | --- | --- | --- |
| 2019 | 70,919 | 1,988,742 | 23,203 (1.17 %) |
| COVID-1 | 485 | 17,756 | 141 (0.79 %) |
| COVID-2 | 1,730 | 59,956 | 460 (0.77 %) |

### A.5 Features used for training models

Patient features are sampled at irregular time intervals reflecting ad hoc clinical measurements taken by hospital staff. Each sample is characterised by a 77-dimensional feature vector and a binary label (retrospectively generated) signifying respiratory deterioration within the next 24 hours. Features include demographic characteristics, vital sign measurements, laboratory test results, and inspired oxygen concentration (FiO_2_). Table 5 documents all features characterising a sample.

**Table 5:**
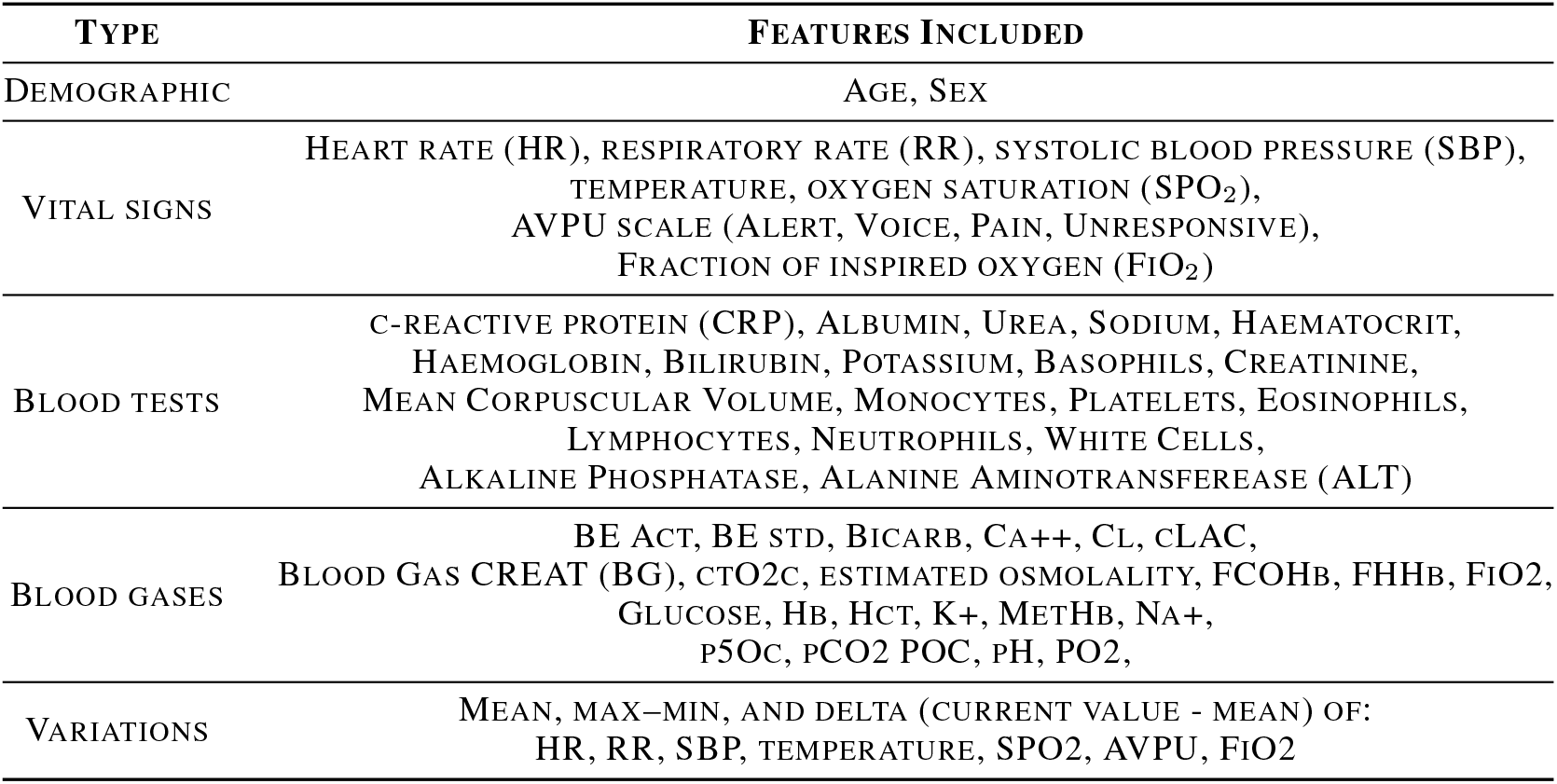
Features used for the task of respiratory deterioration prediction.

### A.6 Distribution shift between 2019 and COVID-1 datasets

Figure 3 highlighting distribution shifts between *2019* and *COVID-1* dataset.

### A.7 Data Availability

The data analysed is not publicly available as it contains personal/sensitive patient information. However, it can be obtained from the Infections in Oxfordshire Research Database (https://oxfordbrc.nihr.ac.uk/research-themes/ modernising-medical-microbiology-and-big-infection-diagnostics/ infections-in-oxfordshire-research-database-iord/), subject to an application and research proposal meeting on the ethical and governance requirements of the database.

## B Model Architecture and Parameter setting

We have used the following neural architecture in this work:

DNN WITH 308 NODES → RELU ACTIVATION → DROPOUT WITH 0.25 RATE → DNN WITH 231 NODES → RELU ACTIVATION → DROPOUT WITH 0.25 RATE → DNN WITH 1 NODE → Sigmoid activation.

The models are trained using Adam optimiser with a fixed learning rate of 10^*−*4^ and a batch size of 2048.

For implementing both GDumb and GEM, we maintain a buffer of 5000 examples (2500 from each class) from the previous domain.

## C Mode/Domain Identification

Suppose we have obtained the incremental mode network as per the proposed method. This network contains *d* modes or domains represented as *d* ∈ ℳ. Then, *θ*_*d*_ and 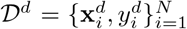 is the mode and the corresponding dataset for domain *d*.

We sample *K* training examples of every class *c* ∈ 𝒞 from each dataset 𝒟^*d*^ and compute mean of penultimate layer embedding for these *K* examples as:

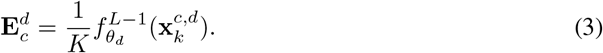

Here, 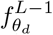 represents the all but the last layer of the model for domain *d*. 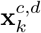 is the *k*th example of *c*th class sampled from 𝒟_*d*_.

For a test example **x**_*t*_, we determine its mode or domain as:

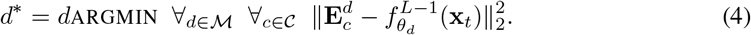

Thus, the embedding of the test example and the chosen *domain representational embedding* (classspecific means) exhibit maximum similarity.

## Footnotes

1 http://github.com/timgaripov/dnn-mode-connectivity

## References

Jacob Armstrong and D Clifton. Continual learning of longitudinal health records. arXiv preprint arXiv:2112.11944, 2021.

Zhiyuan Chen and Bing Liu. Lifelong Machine Learning, chapter 1 & 2. Synthesis Lectures on Artificial Intelligence and Machine Learning. Morgan & Claypool, 2 edition, 2018. ISBN 1681733021; 9781681733029.

Timur Garipov, Pavel Izmailov, Dmitrii Podoprikhin, Dmitry P Vetrov, and Andrew G Wilson. Loss surfaces, mode connectivity, and fast ensembling of dnns. Advances in Neural Information Pro-cessing Systems, 31, 2018.

Cecilia S Lee and Aaron Y Lee. Clinical applications of continual learning machine learning. The Lancet Digital Health, 2(6):e279–e281, 2020.

Xiaoxuan Liu, Livia Faes, Aditya U Kale, Siegfried K Wagner, Dun Jack Fu, Alice Bruynseels, Thushika Mahendiran, Gabriella Moraes, Mohith Shamdas, Christoph Kern, et al. A comparison of deep learning performance against health-care professionals in detecting diseases from medical imaging: a systematic review and meta-analysis. The Lancet Digital Health, 1(6):e271–e297, 2019.

David Lopez-Paz and Marc’Aurelio Ranzato. Gradient episodic memory for continual learning. Advances in neural information processing systems, 30, 2017.

Ameya Prabhu, Philip HS Torr, and Puneet K Dokania. GDumb: A simple approach that questions our progress in continual learning. Proceedings of European Conference on Computer vVision, pp. 524–540, 2020.

Alvin Rajkomar, Eyal Oren, Kai Chen, Andrew M Dai, Nissan Hajaj, Michaela Hardt, Peter J Liu, Xiaobing Liu, Jake Marcus, Mimi Sun, et al. Scalable and accurate deep learning with electronic health records. NPJ digital medicine, 1(1):1–10, 2018.

Haoqi Sun, Eyal Kimchi, Oluwaseun Akeju, Sunil B Nagaraj, Lauren M McClain, David W Zhou, Emily Boyle, Wei-Long Zheng, Wendong Ge, and M Brandon Westover. Automated tracking of level of consciousness and delirium in critical illness using deep learning. NPJ digital medicine, 2(1):1–8, 2019.

Norman Tatro, Pin-Yu Chen, Payel Das, Igor Melnyk, Prasanna Sattigeri, and Rongjie Lai. Optimizing mode connectivity via neuron alignment. Advances in Neural Information Processing Systems, 33:15300–15311, 2020.

Anshul Thakur, Jacob Armstrong, Alexey Youssef, David Eyre, and David A. Clifton. Self-aware sgd: Reliable incremental adaptation framework for clinical ai models. IEEE Journal of Biomedical and Health Informatics, 27(3):1624–1634, 2023. doi: 10.1109/JBHI.2023.3237592.

Jack Wilkinson, Kellyn F Arnold, Eleanor J Murray, Maarten van Smeden, Kareem Carr, Rachel Sippy, Marc de Kamps, Andrew Beam, Stefan Konigorski, Christoph Lippert, et al. Time to reality check the promises of machine learning-powered precision medicine. The Lancet Digital Health, 2(12):e677–e680, 2020.

Alexey Youssef, Samaneh Kouchaki, Farah Shamout, Jacob Armstrong, Rasheed El-Bouri, Thomas Taylor, Drew Birrenkott, Baptiste Vasey, Andrew Soltan, Tingting Zhu, et al. Development and validation of early warning score systems for COVID-19 patients. medRxiv, 2020.

